# Real-world effectiveness of non-pharmaceutical interventions in containing COVID-19 pandemic after the roll-out of coronavirus vaccines: A systematic review

**DOI:** 10.1101/2023.11.07.23297704

**Authors:** Xiaona He, Huiting Chen, Xinyu Zhu, Wei Gao

**Affiliations:** Department of Epidemiology and Health Statistics, School of Public Health, Nanchang University, Nanchang, China; Jiangxi Provincial Key Laboratory of Preventive Medicine, Nanchang University, Nanchang 330006, PR China

**Keywords:** Non-pharmaceutical interventions, Real-world effectiveness, COVID-19, Vaccination

## Abstract

**Background:** Non-pharmaceutical interventions (NPIs) have been widely used to control the transmission of infectious diseases. However, the current research evidence on the policy mechanisms of NPIs is still limited. This study aims to systematically identify, describe, and evaluate the existing literature for the real-world effectiveness of NPIs in containing COVID-19 pandemic after the roll-out of coronavirus vaccines, in order to search for optimal strategies for implementing NPIs.

**Methods:** We conducted a comprehensive search of relevant studies from January 1, 2021, to June 4, 2023 in PubMed, Embase, Web of science and MedRxiv. Two authors independently assessed eligibility and extracted data. Risk of bias assessment tool was used to evaluate the study design, statistical methodology, and quality of reporting. Data were collected, synthesised and analyzed through quantitative and qualitative approaches. The findings were presented using summary tables and figures, including information on the target countries and regions of the study, types of NPIs, and evidence quality.

**Results:** The review included a total of seventeen studies that examined the real-world effectiveness of NPIs in containing the COVID-19 pandemic after the vaccine roll-out. These studies used five composite indicator that combined multiple NPIs and fourteen individual NPIs. The studies had an average quality assessment score of 13 (range: 10-16), indicating moderately high quality. Among the included studies, nine assessed the effectiveness of the composite indicator, with four of them also evaluating individual NPIs. Additionally, twelve studies investigated the effectiveness of individual NPIs. The most frequently evaluated individual NPIs were testing policy, restrictions on gathering, facial covering, and school closure. Workplace closures and stay-at-home requirements were also assessed. The effectiveness of NPIs varied depending on time frames, countries and regions.

**Conclusion:** In summary, the research evidence suggests that NPIs remain effective in curbing the spread of COVID-19 even after the roll-out of vaccines. Studies based on different contexts had different viewpoints or conclusions regarding the effectiveness of NPIs in containing the COVID-19 pandemic. Further research is needed to understand the policy mechanisms and address potential future challenges.

## Background

Since the availability of COVID-19 vaccines, governments worldwide have implemented vaccination and non-pharmaceutical interventions (NPIs) such as testing policies, gathering restrictions, facial covering policies, school closures, workplace closures to contain local transmission of COVID-19[1, 2]. The NPIs, also known as public health measures, aim to break infection chains by altering key aspects of our behavior. Extensive research has been dedicated to examining the effectiveness of NPIs in controlling the outbreak of COVID-19[3–5].

Before the COVID-19 pandemic, there existed literature in addressing the effect of NPI implementation on influenza pandemic[6]. However, a key challenge in this topic is the limited evidence regarding the effectiveness of NPIs, which predominantly relies on mathematical modeling with a limited number of empirical studies[7–9].

Considering the potential harm posed by respiratory infectious disease outbreaks and the high social and economic costs associated with implementing various NPIs, it is essential to conduct research that examines the effectiveness of NPIs in controlling pandemics in real-world settings. Mendez et al. conducted a systematic review and identified that school closures, workplace closures, business and venue shutdowns, and public event restrictions as the most effective measures in controlling the real-world spread of COVID-19[7].

However, various countries implemented diverse NPIs at different stages of the pandemic to control the spread of COVID-19, especially after the introduction of coronavirus vaccines. Asian countries consistently enforced strict NPIs throughout the first half of 2021[10], while no NPIs were implemented in France after May 2021[11]. At the early stage of vaccine roll-out, vaccination coverage in most countries remained relatively low[2]. As of June 30, 2021, a total of 29.29% of the world’s population had received at least one dose of the vaccine, with significant variations in vaccination coverage across countries[2]. Despite an increase in vaccination rates in many countries during the latter half of 2021, the number of confirmed new COVID-19 cases remained high worldwide due to the prevalence of the highly transmissible and immune-escape Delta variant in the second half of 2021[12], followed by the emergence of the Omicron variant in early 2022[13].

The effectiveness of NPIs in controlling the COVID-19 pandemic after the roll-out of vaccines has also received considerable attention[14]. Nevertheless, the policy mechanisms underlying their effectiveness, such as determining when to implement stricter lockdown measures or when to ease restrictions, as well as identifying which types of NPIs are more suitable for different stages, remain unclear.

This review focuses on investigating the real-world effectiveness of NPIs in containing the COVID-19 pandemic after vaccine roll-out. We summarize the current evidence from the real world on the effectiveness, aiming to deepen the current understanding, fill in the gaps in the topics, and provide evidence for the future.

## Methods

The reporting of this review was guided by the Preferred Reporting Items for Systematic Reviews and Meta-Analyses (PRISMA) statement[15]. This review was registered at the international prospective register of systematic reviews (PROSPERO; CRD42023411560).

### Data sources and searches

We conducted a comprehensive search of relevant literature in Embase, PubMed, and Web of Science, and preprints on MedRxiv from January 1, 2021 to June 4 2023. Our search was limited to articles written in English. The search terms included NPIs, COVID-19 and vaccination, which were detailed in Table S1 of the supplemental file. We used EndNote(version 20.0) software to process and remove duplicates. In addition, we manually searched for citations and related articles of the included studies using Google Scholar.

### Study selection and eligibility criteria

One author (XH) screened eligible studies by reviewing the titles and/or abstracts of searched articles. If an article was deemed relevant or if the information provided in the title or abstract was insufficient to make a decision, the full texts were retrieved and examined. For all eligible studies(n=182), two independent authors (XH and HC) assessed the eligibility criteria for each study by evaluating the full text and determining inclusion or exclusion. Any discrepancies between authors were resolved by discussions with the third reviewer (XZ) and the senior author (WG) to reach a consensus.

In general, we adopted an inclusive approach by retaining all studies that could not be excluded with high confidence. All decisions were documented in a spreadsheet. Studies were included in the review if they: 1) assessed the effectiveness of NPIs during the roll-out of COVID-19 vaccines; 2) evaluated the effectiveness of NPIs and vaccination coverage using real-world data; 3) analyzed the respective/interactive impact of NPIs and vaccination coverage; 4) assessed the effectiveness at least one type of NPIs; 5) measured at least one health outcome; 6) obtained evidence through ecological study. Studies were excluded from the review if they: 1) were based on forecasts or simulations; 2) analyzed the impact of adherence or compliance to NPIs and intention or willingness to vaccination; 3) assessed NPI effectiveness in controlling other diseases; 4) did not directly assess the effectiveness of NPIs.

### Quality assessment

To assess the quality of studies, we used a risk of bias assessment tool based on a bibliometric review of ecological studies, as proposed by Dufault et al (2011)[16]. This tool has been previously used and adapted in recent reviews[7, 17, 18]. The purpose of the risk of bias assessment tool is to critically evaluate study design, statistical methodology and practices, and the quality of reporting. Two independent reviewers (XH and HC) evaluated the risk of bias for each included study. Any discrepancies between the reviewers were resolved by discussion with a third reviewer (XZ) and the senior author (WG) to reach a consensus. The checklist of risk of bias assessment tool was included in the Table S2 of Supplemental File.

### Data synthesis and analysis

Characteristics and outcomes of individual studies were extracted, including study authors, year, setting, study design, duration of study, type and/or intensity of NPIs, vaccination coverage, assessment indicators of outcome such as effective reproductive number (Rt), basic reproduction number(R0) and the number of daily new cases or deaths. The classification and intensity of NPIs were mainly based on information from a global panel database of pandemic policies (Oxford COVID-19 Government Response Tracker) [19].

In the final stage of the systematic review, we synthesised the findings from all eligible ecological studies(n=17) to determine the real-world effectiveness of NPIs in containing the COVID-19 pandemic after the vaccine roll-out. Data were collected, synthesised and analyzed using quantitative and qualitative approaches. The results were presented using summary tables and figures, including the target countries and regions of the studies, NPIs types, evidence quality.

## Results

### Summary of literature screening and background

Seventeen ecological studies were included in the review, of which fourteen were published and three were preprints. The PRISMA diagram flow is presented in Figure 1. For more information on excluded articles and reasons for their exclusion, please refer to Table S3 in the Supplemental File. These studies encompass research samples from over 88% of countries and regions worldwide, with each study focusing on a different geographical scope. Table 1 provides a breakdown of the studies: eight evaluated the effectiveness of NPIs in containing the COVID-19 pandemic on a global scale[20–27], three focused on Europe[28–30], two on the United States[31, 32], one on Asia[10], and one each on India[33], France[11] and Korea[34].

**Figure 1.**
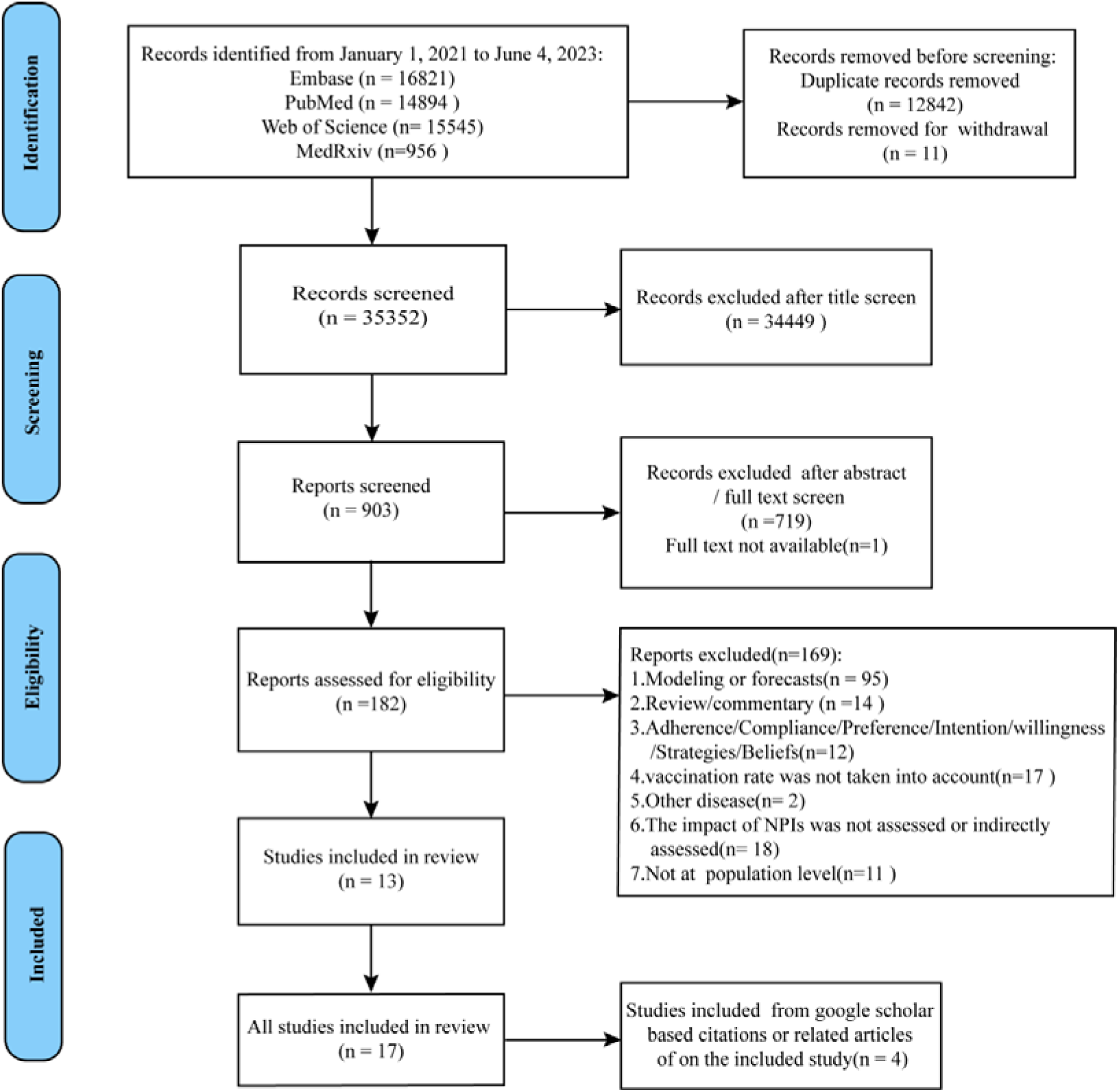
PRISMA flow diagram for the selection of studies.

The seventeen studies examined the impact of NPIs on the COVID-19 pandemic during different periods. eight studies evaluated their effectiveness during the early stage of vaccine roll-out (before July 2021), five during the later stage (the second half of 2021), and the remaining four during the Omicron stage (contain 2022 period).

**Table 1.** Background of included studies.

| Geographical scope | Number of studies | Study time |  |  |
| --- | --- | --- | --- | --- |
|  |  | Early stage | Later stage | Omicron stage |
|  |  | (First half of 2021) | (Second half of 2021) |  |
| worldwide | 8 | 3 | 1 | 4 |
| Europe-wide | 3 | 1 | 2 | 0 |
| US | 2 | 2 | 0 | 0 |
| Asia-wide | 1 | 1 | 0 | 0 |
| India | 1 | 0 | 1 | 0 |
| France | 1 | 1 | 0 | 0 |
| South Korea | 1 | 0 | 1 | 0 |
| Total | 17 | 8 | 5 | 4 |

In terms of quality assessment, the seventeen studies received a moderately high score, averaging 13 (range: 10-16) out of a maximum score of 17. This score reflects the strength of the evidence. The primary sources of risk of bias were the quality of reporting, validity of regression, control of covariates, and internal validity of the methodology. Detailed assessment records can be found in Table S4. Studies used different statistical methods and outcomes, and conducted sensitivity analyses. Only a few studies considered the impact of seasonality.

### Study characteristics

Researchers examined various types of NPIs in the seventeen identified studies. Nine studies evaluated the overall effectiveness of a composite indicator of NPIs (Table 2), while twelve studies specifically assessed the effectiveness of individual NPIs (Table 3). Ge et al., Bollyky et al., and Li et al. analyzed both the effectiveness of the composite indicator of NPIs and individual NPIs. More detailed information can be found in Supplemental File(Table S5).

**Table 2.** Characteristics of studies evaluating the overall effectiveness of a composite indicator of NPIs.

| Author | Publication status, year | Geographical scope | NPIs | Outcome(s) | Score |
| --- | --- | --- | --- | --- | --- |
| Ge et al. (1) | Published, 2022 | European, 33 countries | Stringency index<sup>a</sup> | Rt | 16 |
| Bollyky et al. | Published, 2023 | US, 52 states | Policy mandates<sup>b</sup> | Cases, deaths | 16 |
| Zhou et al. | Published, 2022 | European, 22 countries | Stringency index | Cases, deaths, excess mortality | 15 |
| Paireau et al. | Published, 2023 | France, 96 departments | Lockdown<sup>c</sup> | Rt | 15 |
| Caixia Wang et al. | Published, 2023 | Worldwide, 176 countries | Stringency index | Deaths | 14 |
| Li et al. | Published, 2022 | Worldwide, 8 countries | Combination of four NPIs<sup>d</sup> | Rt | 14 |
| Hale et al. | Published, 2021 | Worldwide, 10 countries | Stringency index | Deaths | 13 |
| Hongjian Wang et al. | Published, 2023 | Worldwide, 176 countries | Stringency index | R0 | 11 |
| Kijin Kim et al. | Published, 2023 | Korea | Social distancing policy index<sup>e</sup> | Cases | 10 |

Notes: a= stringency index included eight containment and closure, including school closure, workplace closure, cancel public events, restrictions on gathering size, close public transport, stay-at-home requirements, restrictions on internal movement, restrictions on international travel ; b =a summary measure that captures a state's use of physical distancing and mask mandates ; c=a comprehensive measure captures the level of social distancing; d=the combination of school closure, workplace closure, restrictions on mass gatherings and stay-at-home requirements; e=restrictiveness of government containment and closure measures implemented; Rt=effective reproductive number; R0=basic reproduction number.

**Table 3.**
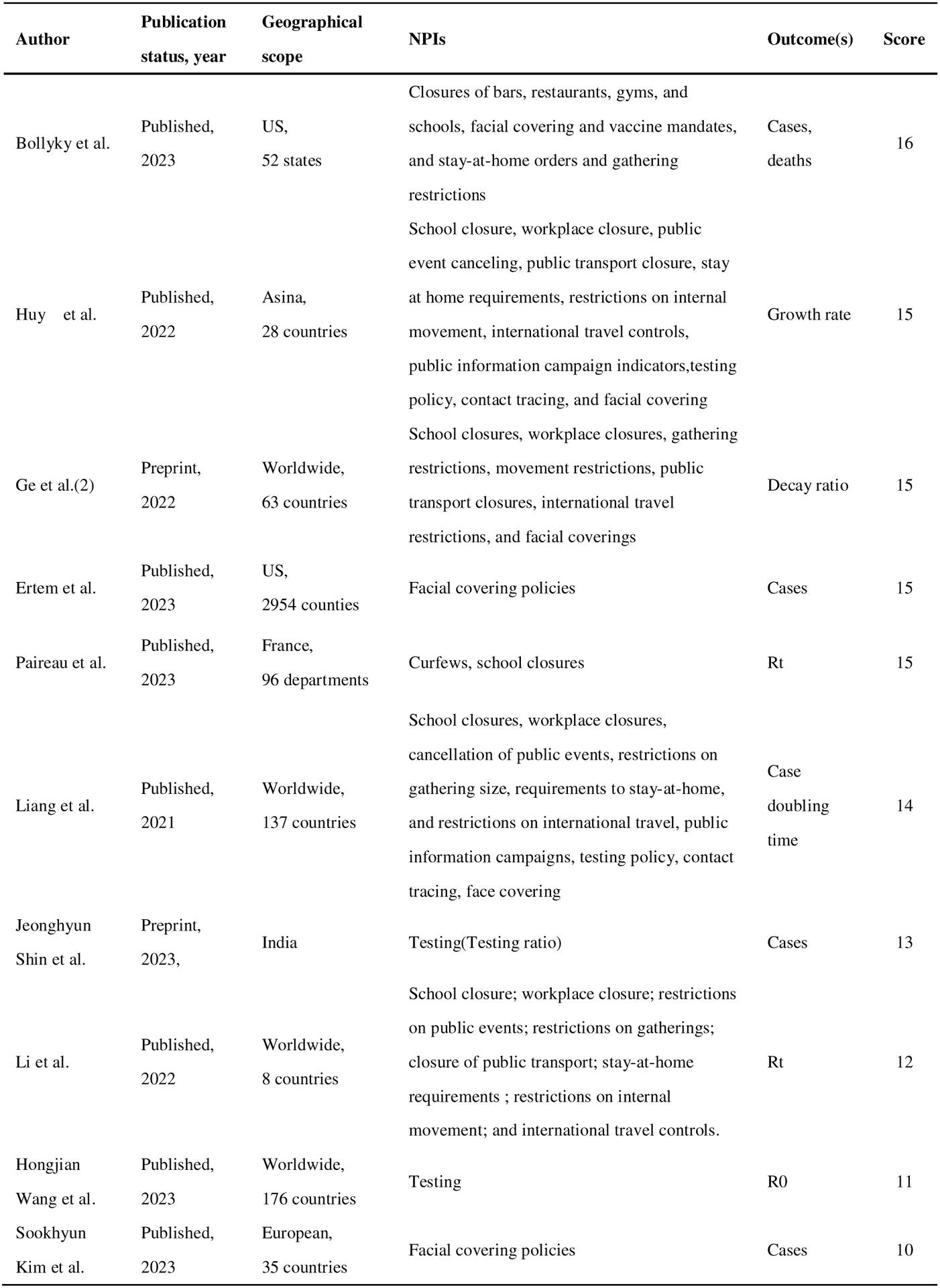
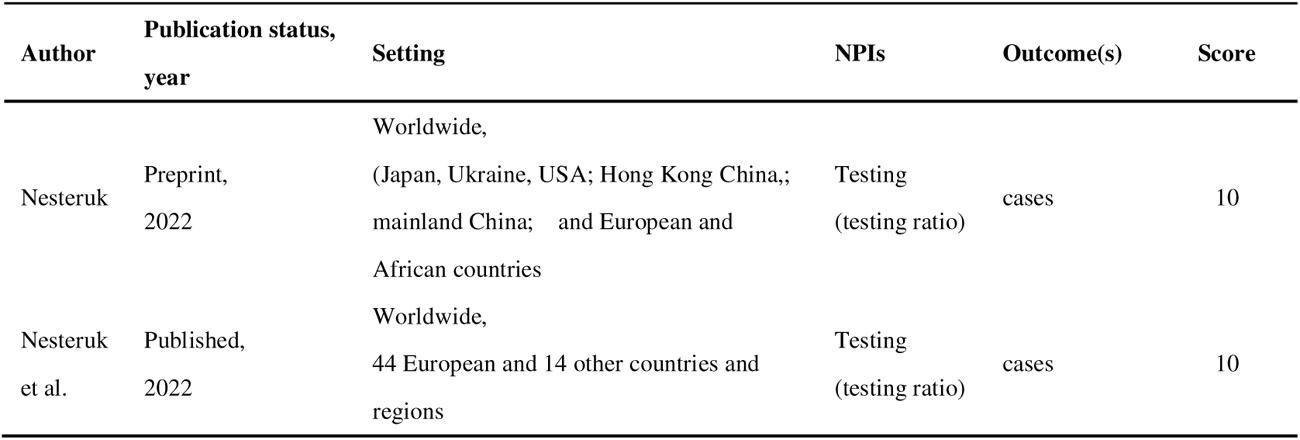
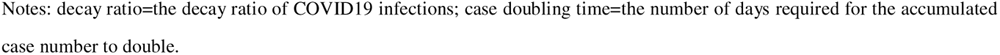
Characteristics of studies evaluating the effectiveness of individual NPIs.

| Author | Publication status, year | Geographical scope | NPIs | Outcome(s) | Score |
| --- | --- | --- | --- | --- | --- |
| Bollyky et al. | Published, 2023 | US, 52 states | Closures of bars, restaurants, gyms, and schools, facial covering and vaccine mandates, and stay-at-home orders and gathering restrictions | Cases, deaths | 16 |
| Huy et al. | Published, 2022 | Asina, 28 countries | School closure, workplace closure, public event canceling, public transport closure, stay at home requirements, restrictions on internal movement, international travel controls, public information campaign indicators, testing policy, contact tracing, and facial covering | Growth rate | 15 |
| Ge et al.(2) | Preprint, 2022 | Worldwide, 63 countries | School closures, workplace closures, gathering restrictions, movement restrictions, public transport closures, international travel restrictions, and facial coverings | Decay ratio | 15 |
| Ertem et al. | Published, 2023 | US, 2954 counties | Facial covering policies | Cases | 15 |
| Paireau et al. | Published, 2023 | France, 96 departments | Curfews, school closures | Rt | 15 |
| Liang et al. | Published, 2021 | Worldwide, 137 countries | School closures, workplace closures, cancellation of public events, restrictions on gathering size, requirements to stay-at-home, and restrictions on international travel, public information campaigns, testing policy, contact tracing, face covering | Case doubling time | 14 |
| Jeonghyun Shin et al. | Preprint, 2023, | India | Testing(Testing ratio) | Cases | 13 |
| Li et al. | Published, 2022 | Worldwide, 8 countries | School closure; workplace closure; restrictions on public events; restrictions on gatherings; closure of public transport; stay-at-home requirements ; restrictions on internal movement; and international travel controls. | Rt | 12 |
| Hongjian Wang et al. | Published, 2023 | Worldwide, 176 countries | Testing | R0 | 11 |
| Sookhyun Kim et al. | Published, 2023 | European, 35 countries | Facial covering policies | Cases | 10 |

Notes: decay ratio=the decay ratio of COVID19 infections; case doubling time=the number of days required for the accumulated case number to double.

Table 3 Continued
| Author | Publication status,<br>year | Setting | NPIs | Outcome(s) | Score |
| --- | --- | --- | --- | --- | --- |
| Nesteruk | Preprint,<br>2022 | Worldwide,<br>(Japan, Ukraine, USA; Hong Kong China,;<br>mainland China; and European and<br>African countries | Testing<br>(testing ratio) | cases | 10 |
| Nesteruk<br>et al. | Published,<br>2022 | Worldwide,<br>44 European and 14 other countries and<br>regions | Testing<br>(testing ratio) | cases | 10 |

The composite indicator of NPIs in this review primarily included five types: stringency index, policy mandates, social distancing policy index, lockdown, and combination of four NPIs(school closure, workplace closure, restrictions on mass gatherings, and stay-at-home requirements). The specific names for these measures varied depending on their sources. The data on NPIs in these studies mainly came from OxCGRT, with additional sources including The Yale State and Local COVID-19 restriction database, governmental websites, and others. These composite indicators were calculated by combining multiple containment and closure measures, representing the overall intensity of various containment and closure policies to some extent. The individual NPIs included containment and closure measures, as well as health systems indicators. Containment and closure measures primarily encompassed restrictions on gatherings, school closures, workplace closures, and stay-at-home requirements. The evaluated health systems indicators included testing policy, facial coverings, contact tracing, public information campaigns, and vaccine mandates.

The studies on vaccination included data on the administration of the first dose, full vaccination, and booster doses according to the vaccination protocol. Two studies did not explicitly specify the doses. The outcome assessed mainly included cases, deaths, instantaneous reproduction number (Rt), and others.

### The effectiveness of composite indicator of NPIs for containing the COVID-19 pandemic after the roll-out of COVID-19 vaccines

As shown in Table 4, even after the introduction of vaccines, the implementation of containment and closure measures continued to be regarded as effective in curtailing the spread of COVID-19. Some studies also compared the effectiveness of NPIs during that period with the impact of vaccination coverage in mitigating the transmission of COVID-19 within populations. Specifically, NPIs were found to be more effective than vaccination in mitigating the spread of COVID-19 during early stage of the vaccination implementation. In the latter half of 2021, the effectiveness of NPIs had relatively diminished compared to the earlier stages. During the Omicron stage, the measures implemented to control the spread of COVID-19 were more effective than vaccination coverage.

In addition to assessing the effectiveness of combinations of containment and closure measures in controlling the spread of COVID-19(cases and Rt), NPIs were also considered in reducing the number of COVID-19 related deaths. Bollyky et al.’s study did not find evidence supporting the effectiveness of NPIs in reducing COVID-19 related deaths, while other studies (Zhou et al., Hale et al. and Caixia Wang et al.) suggested that NPIs could reduced the number of deaths.

**Table 4.** The effectiveness of a composite indicator of NPIs in the studies.

| Stage | Author | Geographical scope | NPIs assessed | Outcome | Effectiveness | Compare to vaccination coverage | Score |
| --- | --- | --- | --- | --- | --- | --- | --- |
| Early | Ge et al.(1) | European | Stringency index | cases | effective | higher | 16 |
| Early | Bollyky et al. | United States | Policy mandates | cases | effective | not compared | 16 |
| Early | Zhou et al. | European | Stringency index | cases | effective | higher | 15 |
| Early | Paureau et al. | France | Lockdown | Rt | effective | higher | 15 |
| Early | Zhou et al. | European | Stringency index | deaths | effective | not compared | 15 |
| Early | Hale et al | Worldwide | Stringency index | deaths | effective | not compared | 13 |
| Later | Ge et al.(1) | European | Stringency index | cases | effective | lower | 16 |
| Later | Li et al. | Worldwide | Combination of four NPIs | cases | effective | not compared | 14 |
| Later | Kijin Kim et al. | South Korea | Social distancing index | cases | effective | lower | 10 |
| Omicron | Hongjian Wang et al. | Worldwide | Stringency index | cases | effective | higher | 11 |
| Omicron | Bollyky et al. | United States | Policy mandates | deaths | not effect | not compared | 16 |
| Omicron | Caixia Wang et al. | Worldwide | Stringency index | deaths | Effective | not compared | 14 |

### The relative effectiveness of individual NPIs for containing the COVID-19 pandemic after the roll-out of COVID-19 vaccines

Twelve studies have evaluated the individual effects of NPIs on controlling the COVID-19 pandemic after the roll-out of COVID-19 vaccines. A total of 14 individual NPIs were assessed in this review. Among the<u>m</u>, testing policies, restrictions on gatherings, facial coverings, and school closures were mentioned most frequently. Additionally, workplace closures, stay-at-home requirements, and restrictions on international travel were also highlighted. Figure 2 demonstrates these findings.

Moreover, based on a compilation of the evidence from the included twelve studies, testing policies(4/6), workplace closures(3/5), and restrictions on gatherings(3/6) were considered the most effective NPIs in containing the COVID-19 pandemic following the roll-out of vaccines.

**Figure 2.**
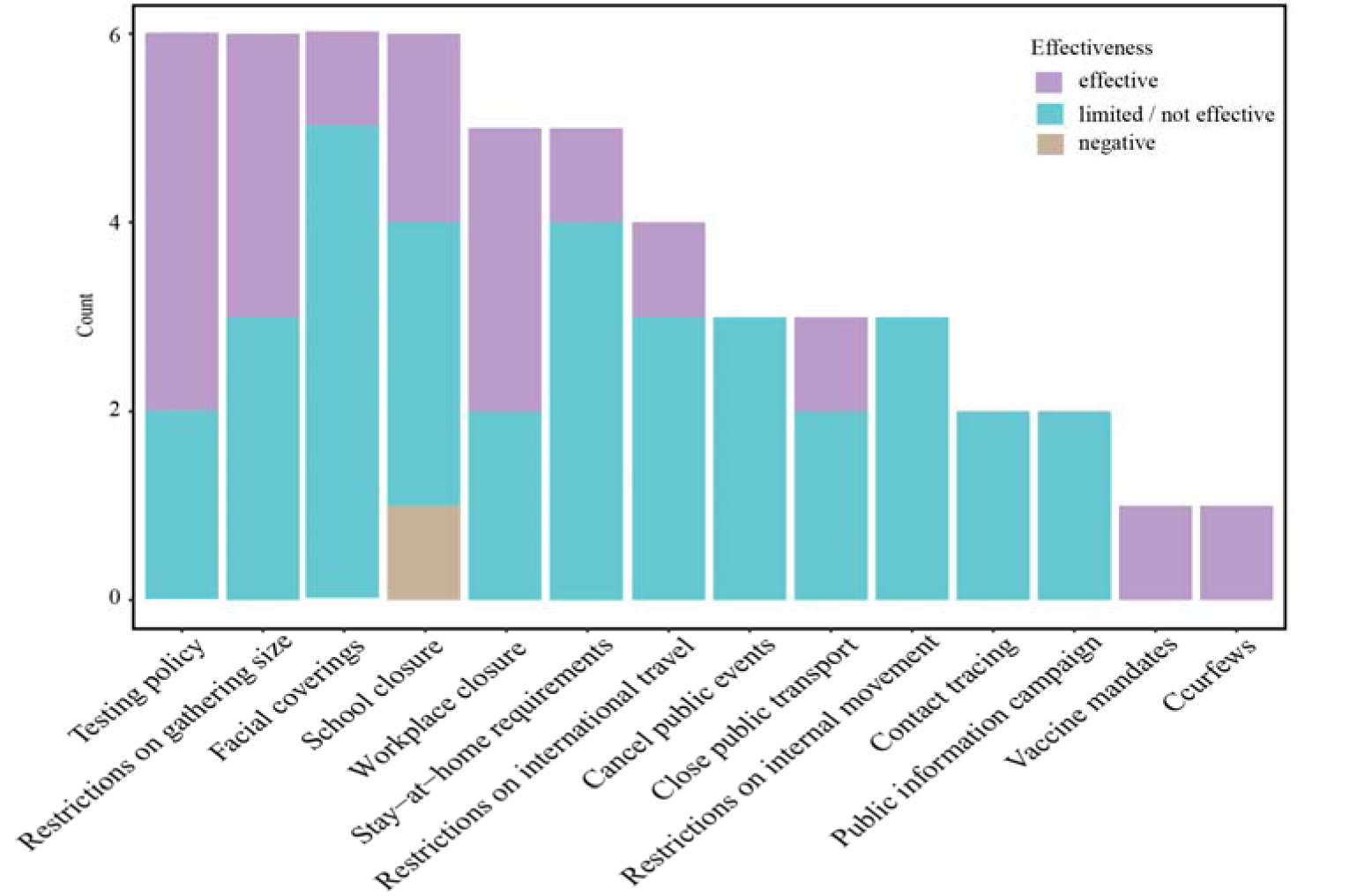
The relative effectiveness of individual NPIs for containing the COVID-19 pandemic after the roll-out of COVID-19 vaccines. The Y-axis represents the count of assessments for this NPIs. The colors of the stacked bar represent the effectiveness of assessed NPIs in containing the COVID-19 pandemic following the roll-out of vaccines. The purple color indicates that a study considers the NPI to be the most effective measure. The blue color signifies limited effectiveness or a lack of association with containing the COVID-19 pandemic, according to the study. The brown color represents a negative correlation between the NPI and containing the spread of COVID-19.

We categorised the included studies according to the target countries and regions, as shown in Table 5. The types of NPIs evaluated and the effective NPIs identified varied across different geographical locations. Based on global studies, testing policy, restrictions on gathering size, workplace closure, school closure, stay-at-home requirements, and restrictions on international travel were found to be relatively effective. In Asian studies, restrictions on gathering size and the closure of public transport were considered as effective measures. In studies conducted in the United States, only vaccination mandates were deemed effective. Data from France indicated that curfews were effective. Face covering mandates were associated with a decrease in COVID-19 incidence in European countries. Testing was effective in India during the vaccination stage.

**Table 5.** The types and the relative effective individual NPIs were classified according to the study’s target countries and regions.

| Author | Geographical scope | NPIs assessed | Effective intervention measures | Score |
| --- | --- | --- | --- | --- |
| Huy et al. | Asia | School closure, workplace closure, cancel public events, restrictions on gathering size, close public transport, stay-at-home requirements, restrictions on internal movement, restrictions on international travel, public information campaign, testing policy, contact tracing, facial coverings | Restrictions on gathering size, Close public transport | 15 |
| Liang et al. | Worldwide | School closure, workplace closure, cancel public events, restrictions on gathering size, stay-at-home requirements, restrictions on international travel, public information campaign, testing policy, contact tracing, facial coverings | School closure, workplace closure | 14 |
| Li et al. | Worldwide | School closure, workplace closure, cancel public events, restrictions on gathering size, close public transport, stay-at-home requirements, restrictions on internal movement, restrictions on international travel | School closure, workplace closure, restrictions on gathering size, stay-at-home requirements | 12 |
| Ge et al.(2) | Worldwide | School closure, workplace closure, restrictions on gathering size, close public transport, restrictions on international travel, facial coverings, restrictions on internal movement, stay-at-home requirements | Workplace closure, restrictions on gathering size, restrictions on international travel | 15 |
| Nesteruk | Worldwide | Testing policy | Testing policy | 10 |
| Nesteruk et al. | Worldwide | Testing policy | Testing policy | 10 |
| Hongjian Wang et al. | Worldwide | Testing policy | Testing policy | 11 |
| Bollyky et al. | US | Workplace closure, School closure, Restrictions on gathering size, Facial coverings, Stay-at-home requirements, Vaccine mandates, Restrictions on gathering size | Vaccine mandates | 16 |
| Ertem et al. | US | Facial coverings | None | 15 |
| Paureau et al. | France | Curfews, school closure | Curfews | 15 |
| Sookhyun Kim et al. | European | Facial coverings | Facial coverings | 10 |
| Shin et al. | India | Testing policy | Testing policy | 13 |

## Discussion

### Summary of the main findings

The types of NPIs evaluated and the effective NPIs identified varied across different periods and geographical locations. Overall, our research shows that NPIs continued to be effective in controlling the spread of COVID-19 even after the roll-out of vaccines. Our previous research work also supports this conclusion[35]. The most frequently evaluated NPIs included testing policies, restrictions on gatherings, facial coverings, and school closures, followed by workplace closures, stay-at-home requirements, and restrictions on international travel.

The overall effectiveness of a composite indicator of NPIs varied depending on the period, with factors such as the intensity of implementation, compliance, increasing vaccine coverage, and the emergence of VOCs playing a role. NPIs remained important for mitigating the pandemic in the early stage of the vaccination when coverage was low[11, 28, 30]. However, as vaccine coverage increased, their marginal effects were surpassed by vaccination[28, 34]. In Omicron stage, measures were more effective in controlling the spread of COVID-19 than vaccination coverage due to the high immune evasion capability of the Omicron variant[13, 27]. It is important to note that NPIs and vaccinations work through different mechanisms to combat the pandemic[36]. NPIs physically reduce population contact and transmission of the virus, while vaccinations reduce susceptible populations by enhancing immunity. Overall, a combination of both containment and closure measures and vaccination is recommended to contain COVID-19 after after the vaccine has been introduced[11, 22, 27, 28, 30, 34].

The types of the evaluated NPIs, as well as the effective NPIs, varied across different target countries and regions. Factors such as differences in government effectiveness[23], culture[22], and economic disparities among different countries and regions may affect the effectiveness of various NPIs[37].

Testing policies is a central pillar of public health response to global health emergencies. In the included studies, testing policies were primarily evaluated during the Omicron period. Nesteruk[24], Nesteruk et al.[25], Wang et al.[27], and Shin et al.[33] found that strengthening testing could reduce the number of COVID-19 infections. In addition, Shao et al also found that large-scale SARS-CoV-2 rapid antigen testing alleviated the Omicron outbreak in China[38].

Gathering restrictions are primarily implemented to curb the spread of infectious diseases by reducing interpersonal contact, which can occur through various transmission pathways such as droplets, direct contact, and aerosols[39]. A previous study indicated that restrictions on gathering had the greatest contribution (37.60%) to suppressing influenza transmission during the 2019-2020 influenza season[40]. Different levels of gathering restrictions have shown varying effectiveness. According to the categorization by OxCGRT, the levels of restrictions on gatherings range from strictest to the weakest, including limitations on gatherings of 10 or fewer people, 11-100 people, 101-1000 people, and gatherings with 1000 or more individuals[1]. Studies by Huy et al. suggested that limiting gatherings to 10 or fewer people was most strongly correlated with a decrease in COVID-19 case numbers[10]. A similar finding was also supported by research conducted by Liang et al[23].

Facial coverings are a form of personal protective equipment used to shield the face from various external hazards like splashes, droplets, and aerosols. Among the summarized evidence, only one study (Sookhyun Kim et al.)[29] found that the incidence of COVID-19 was significantly higher after the relaxation of face covering mandates. Other studies found no association between the implementation of facial covering policies and a reduction in COVID-19 cases[10, 20, 23, 31, 32]. Facial covering policies do not represent the actual use of masks for preventing infections but rather serve as public health measures. The effectiveness of facial covering policies depends on compliance with the policies, proper mask usage, and the duration of mask-wearing. Bollyky et al. found no evidence that implementing facial covering policies reduced the number of COVID-19 infections, but they did observ an association between mask use and lower rates of COVID-19 infection[31].

The effectiveness of school closures in reducing COVID-19 infections appears to be controversial. Liang et al.[23] and Li et al. [22] argued that school closures were associated with mitigating the spread of COVID-19, while Paireau et al.[11] and Ge et al.[20] suggested that their effectiveness was limited. Conversely, Huy et al.[10] discovered that the policy of school closure had the opposite effect on the reduction of infection rate. A previous study found that the policy had a potential for effectively reducing influenza transmission[41]. However, the optimum strategy of the policy of school closures remains unclear, whether in controlling the spread of influenza or COVID-19 pandemic.

Liang et al.[23], Li et al.[22], and Ge et al[20]. considered workplace closure as an effective intervention in containing the spread of COVID-19, after the roll-out of coronavirus vaccines. Modeling studies estimated that implementing only workplace social distancing measures could reduce the median cumulative incidence of influenza in the general population by 23% from 2000 to 2017[42].

Stay-at-home orders[22], restrictions on international travel[20], public transport closures[10], vaccine mandates[31], and curfews[11] have been identified as effective measures in controlling the spread of COVID-19 according to a minority of included studies after the introduction of vaccines. Additionally, there is no evidence to suggest that restrictions on internal movement[10, 20, 22], public information campaigns[10, 23], and contact tracing[10, 23] were associated with a reduction in the transmission of COVID-19. Considering the number and heterogeneity of existing evidences, further research is needed to identify the impact and mechanisms of the implementation of these NPIs in controlling the spread of COVID-19.

There is controversy surrounding whether NPIs can effectively reduce COVID-19 deaths. Studies have shown that NPIs do not directly reduce the number of COVID-19 deaths[43]. However, research conducted by Hale et al.[21] and Wang et al.[26] found that, a higher stringency index was associated with a lower average daily death toll. It is possible that NPIs indirectly reduce the number of deaths by mitigating the spread of COVID-19. Nevertheless, these studies lack analysis or explanation regarding the specific indirect impacts.

The research on NPIs’ effectiveness in reducing the transmission of infectious diseases, especially respiratory ones like SARS, influenza, and COVID-19, has always received attention. However, our understanding of the effectiveness of these measures in controlling respiratory infectious diseases is still not comprehensive enough, even in the context of the ongoing COVID-19 pandemic, particularly since the introduction of vaccines.

### Strengths and limitations

This study has several strengths. Firstly, we conducted a systematic and comprehensive search across various databases to investigate the real-world effectiveness of NPIs in containing the COVID-19 pandemic post-vaccine roll-out. Secondly, we employed a risk of bias assessment tool to critically assess the potential biases in the included studies. Thirdly, we summarized and analyzed the available evidence using quantitative and qualitative approaches, presenting the findings in tables and figures. Nonetheless, our study also has limitations. Firstly, we included three preprints that had not been peer-reviewed, although we did evaluate their risk of bias. Secondly, due to variations in study design, analytical methodologies, and outcome measures, we were unable to perform a meta-analysis. Lastly, the evidence derived from the included studies was limited as they relied on retrospective and observational data, which cannot establish a causal relationship between NPIs and outcomes due to potential confounding variables.

## Conclusion

In conclusion, the understanding of the effectiveness of NPIs in mitigating the pandemic following vaccination is inadequate. The NPIs had varying effects in containing COVID-19 pandemic in different geographical areas. Future researchers must continue to focus on the effectiveness of different NPIs in various contexts, to ascertain when to ease restrictions and when to strengthen them. It is essential to comprehend the policy mechanisms of these intervention measures in controlling the spread of COVID-19 and other respiratory infectious diseases, such as influenza.

## Data Availability

All data produced in the present study are available upon reasonable request to the authors.

## List of abbreviations

NPIs: Non-pharmaceutical interventions
COVID-19: Coronavirus disease 2019
Rt: Effective reproductive number
R0: Basic reproduction number

## Declarations

### Ethics approval and consent to participate

Not applicable.

### Consent for publication

Not applicable.

### Availability of data and materials

All data were collected from publicly available literatures, and all data generated or analyzed during this study are included in this article and its supplemental files.

### Authors’ contributions

WG conceived the study and devised the methodology. XH performed the literature search, literature screening, data extraction, management and analysis. HC and XZ reviewed the literature and conducted the collection and curation of data. XH drafted the manuscript. WG directed the study, and critically revised the manuscript. All authors had full access to all the data in the study and verified the data. WG had final responsibility for the decision to submit for publication.

## Acknowledgements

Not applicable.

