## Supplemental File TableS1-S5 for "Real-world effectiveness of non-pharmaceutical interventions in containing COVID-19 pandemic after the roll-out of coronavirus vaccines: A systematic review"

Supplementary Appendix

Table S1 Search terms	2

Table S2 Checklist of risk of bias assessment tool	3

Table S3 Excluded articles and reasons for their exclusion	4

Table S4 Quality assessment	7

Table S5 The characteristic of the included studies	8

**Table S1 Search terms**

| **Search terms** | **Terms** |
| --- | --- |
| COVID-19/SARS-CoV-2 | 2019 nCoV Infection |
|  | SARS-CoV-2 Infection |
|  | 2019 Novel Coronavirus Disease |
|  | 2019 Novel Coronavirus Infection |
|  | COVID-19 Virus Infection |
|  | Coronavirus Disease 2019 |
|  | Severe Acute Respiratory Syndrome Coronavirus 2 Infection |
|  | COVID-19 Virus Disease |
|  | SARS Coronavirus 2 Infection |
|  | 2019-nCoV Disease |
|  | COVID-19 Pandemic |
| vaccination | vaccinations |
| NPIs | non-pharmaceutical intervention |
|  | public policy |
|  | lockdown |
|  | school closure |
|  | social distance |
|  | mask |
|  | containment policy |
|  | isolation |
|  | stay at home |
|  | containment measure |
|  | quarantine |
|  | covid-19 testing |
|  | contact tracing |
|  | government response |
|  | public transport |
|  | travel |
|  | gathering |
|  | workplace closure |
|  | public event |
|  | internal movement |
|  | fiscal measures |
|  | international support |
|  | containment measure |
|  | income support |
|  | public information campaigns |
|  | public information campaign |
|  | investment |
|  | aged protection |

**Table S2 Checklist of risk of bias assessment tool**

| **Category** | **Evaluation Criterion** | **Score** |
| --- | --- | --- |
| Study design（Max=7） | Design | Cross-sectional=0；longitudinal=1 |
|  | Source of data（Validity of the sources of data to represent the level that it refers to) | No=0; Yes=1 |
|  | Sample size(Number of ecologic units included in the analysis as proportion of the total number of units) | < 10% units =0 10%-80% units = 1  ≥ 80% units = 2 |
|  | Unbiased inclusion of units (Were the units included representative of the group for which inferences are being drawn? For example, for worldwide inferences, inclusion of only developed countries would be biased ) | No=0; Yes=1 |
|  | Level of inference(whether the inferences were made at the group level or the individual level) | Individual level/unclear=0; Group level=1 |
|  | Prespecification of ecological units(whether the ecologic units were explicitly chosen to suit the hypothesis or were seemingly motivated by convenience or necessity ) | No=0; Yes=1 |
| Statistical methodology (Max=7) | Analytical methodologies | Less sophisticated, less flexible=1; more sophisticated and/or more flexible=2 |
|  | Validity of regression(whether there were more than 10 ecologic units per covariate ) | No=0; Yes=1 |
|  | Spatial effects(Inclusion of spatial analysis) | No=0; Yes=1 |
|  | Use of covariates (Did authors adjust analysis for desirable variables? ) | No=0; Yes=1 |
|  | Proper adjustment for covariates | No=0; Yes=1 |
|  | Internal validity of the methodology(Did the authors perform sensitivity analyses or robustness checks?) | No=0; Yes=1 |
| Quality of reporting（max=3） | Statement of study design(whether the authors mentioned one of the keywords ‘‘ecologic,’’ ‘‘ecological,’’ or ‘‘aggregate’’ in their article, excluding references ) | No=0; Yes=1 |
|  | Justification of study design(whether the authors. explicitly justified an ecologic analysis ) | No=0; Yes=1 |
|  | Discussion cross-level bias and limitations(whether the authors sufficiently cautioned readers, in interpreting their results, against making heedless individual-level inference ) | No=0; Yes=1 |

Note: The risk of bias assessment tool, as described by Dufault et al (2011), originates from their study titled “The quality of modern cross-sectional ecologic studies: a bibliometric review” published in the American Journal of Epidemiology. This tool is used to evaluate the quality and potential bias in cross-sectional ecologic studies.

**Table S3 Excluded articles and reasons for their exclusion**

| **Reasons for their exclusion** | **N** | **Title of excluded articles** |
| --- | --- | --- |
| Modeling or forecasts | 95 | Unraveling the dynamics of the Omicron and Delta variants of the 2019 coronavirus in the presence of vaccination, mask usage, and antiviral treatment  Effect of hybrid immunity, school reopening, and the Omicron variant on the trajectory of the COVID-19 epidemic in India: a modelling study  Modified susceptible–exposed–infectious–recovered model for assessing the effectiveness of non-pharmaceutical interventions during the COVID-19 pandemic in Seoul  Vaccination and three non-pharmaceutical interventions determine the dynamics of COVID-19 in the US  Modeling the Impact of nonpharmaceutical interventions on COVID-19 transmission in K-12 Schools  Early guidance for Sars-Cov-2 health policies in India: Social Distancing amidst Vaccination and Virus Variants  Effect of travel restrictions, contact tracing and vaccination on control of emerging infectious diseases: transmission of COVID-19 as a case study  Impacts of social distancing, rapid antigen test and vaccination on the Omicron outbreak during large temperature variations in Hong Kong: A modelling study  From pandemic to a new normal: Strategies to optimise governmental interventions in Indonesia based on an SVEIQHR-type mathematical model  ... |
| Review/commentary | 14 | COVID-19 exit strategy during vaccine implementation: a balance between social distancing and herd immunity；  Can a combination of vaccination and face mask wearing contain the COVID-19 pandemic?  Non-pharmaceutical interventions during the roll out of covid-19 vaccines；  How will mass-vaccination change COVID-19 lockdown requirements in Australia?  Reopening International Borders without Quarantine: Contact Tracing Integrated Policy against COVID-19；  Estimating data-driven COVID-19 mitigation strategies for safe university reopening；  Despite vaccination, China needs non-pharmaceutical interventions to prevent widespread outbreaks of COVID-19 in 2021；  Importance of non-pharmaceutical interventions in the COVID-19 vaccination era: A case study of the Seychelles；  Barrier gesture relaxation during vaccination campaign in France: modelling impact of waning immunity；  Controlling the pandemic during the SARS-CoV-2 vaccination rollout；  COVID-19: the implications and consequences of prolonged lockdown and COVID-19 vaccine cost in a low-middle income country；  COVID-19 air travel restrictions and vaccine passports: An ongoing debate；  Impact of the Delta variant on vaccine efficacy and response strategies；  Vaccines, masks, distancing and credibility: An urgent warning for pandemic management； |
| Adherence/Compliance/Preference/Intention/willingness/Strategies/Beliefs | 12 | Vaccination under the Pandemic and Political Support  Pros and cons factors influence population attitudes toward non-pharmaceutical interventions and vaccination during post-COVID-19  COVID-19 vaccination strategies and policies in India: The need for further re-evaluation is a pressing priority  Vaccination or restriction?: COVID-19 vaccine hesitancy and vaccine passports  Messaging Strategies for Mitigating COVID-19 Through Vaccination and Nonpharmaceutical Interventions  COVID-19 Health Beliefs Regarding Mask Wearing and Vaccinations on Twitter: Deep Learning Approach  COVID-19 screening, testing and vaccination: Perceptions from emergency medicine residents and medical students  Joint analysis of the intention to vaccinate and to use contact tracing app during the COVID-19 pandemic  Vaccination or NPI? A conjoint analysis of German citizens' preferences in the context of the COVID-19 pandemic  The U.S. COVID-19 Trends and Impact Survey, 2020-2021: Continuous real-time measurement of COVID-19 symptoms, risks, protective behaviors, testing and vaccination  Mask is a must: the need of protection and safety against COVID-19  The obligation to use face masks in public spaces as a public health measure and permissible limits on civil liberties |

**Table S3 Continued**

| **Reasons for their exclusion** | **N** | **Title of excluded articles** |
| --- | --- | --- |
| Vaccination rate was not taken into account | 17 | Data-driven multiscale dynamical framework to control a pandemic evolution with non-pharmaceutical interventions  Estimated Mask Use and Temporal Relationship to COVID-19 Epidemiology of Black Lives Matter Protests in 12 Cities  Estimating the risk reduction of isolation on COVID-19 non-household transmission and severe/critical illness in non-immune individuals: September to November 2021  Effectiveness of social distancing interventions in containing COVID-19 incidence: International evidence using Kalman filter  Transmission Dynamics of COVID-19 in Ghana and the Impact of Public Health Interventions  The impact of non-pharmaceutical interventions on COVID-19 cases in South Australia and Victoria  Assessing the impact of non-pharmaceutical interventions (NPIs) and BCG vaccine cross-protection in the transmission dynamics of SARS-CoV-2 in eastern Africa  COVID-19 pandemic over 2020 (withlockdowns) and 2021 (with vaccinations): similar effects for seasonality and environmental factors  Analysis of the Effectiveness of Measures on the COVID-19 Vaccination Rate in Hong Kong  COVID-19 evolution during the pandemic - Implications of new SARS-CoV-2 variants on disease control and public health policies  Impact of the COVID-19 lockdown on routine vaccination in Pakistan: a hospital-based study  The role of mask mandates, stay at home orders and school closure in curbing the COVID-19 pandemic prior to vaccination  ... |
| Other disease | 2 | Control Influenza in Megacities: An Agent-Based Modeling Study With Large-Scale Trajectory Data  Impact of non-pharmaceutical interventions on the incidences of vaccine-preventable diseases during the COVID-19 pandemic in the eastern of China |
| The impact of NPIs was not assessed or indirectly assessed | 17 | To open or not to open: the moderating effects of human mobility on the relationship between vaccination and COVID-19 transmission  Vaccine Effectiveness, School Reopening, and Risk of Omicron Infection Among Adolescents Aged 12-17 Years  Strengthening Social Compact and Innovative Health Sector Collaborations in Addressing COVID-19 in South African Workplaces  The effectiveness of various control strategies: An insight from a comparison modelling study  COVID-19 prevention and control strategies: learning from the Macau model  The effectiveness of post-vaccination and post-infection protection in the hospital staff of three prague hospitals: A cohort study of 8-month follow-up from the start of the covid-19 vaccination campaign (covaness)  Relationship between the use of nonpharmaceutical interventions and COVID-19 vaccination among U.S. child care providers: A prospective cohort study  COVID-19 vaccination roll-outs in small countries within the European region; lessons from eleven countries in the World Health Organization's Small Countries Initiative  Easing Restrictions During Vaccine Scarcity. How Mitigation Measures Help Tackling Associated Moral and Behavioral Challenges  The new UK SARS-CoV-2 variant and lockdown - Causes and consequences  Implications of COVID-19 vaccination and public health countermeasures on SARS-CoV-2 variants of concern in Canada: evidence from a spatial hierarchical cluster analysis  Comparative transmission of SARS-CoV-2 Omicron (B.1.1.529) and Delta (B.1.617.2) variants and the impact of vaccination: national cohort study, England.  Evaluating the effectiveness of lockdowns and restrictions during SARS-CoV-2 variant waves in the Canadian province of Nova Scotia.  Case clustering, contact stratification, and transmission heterogeneity of SARS-CoV-2 Omicron BA.5 variants in Urumqi, China: An observational study  ... |

**Table S3 Continued**

| **Reasons for their exclusion** | **N** | **Title of excluded articles** |
| --- | --- | --- |
| Not at population level | 11 | Association between COVID-19 and consistent mask wearing during contact with others outside the household-A nested case-control analysis, November 2020-October 2021  The Effect of Preventive Measures and Vaccination against SARS-CoV-2 on the Infection Risk, Treatment, and Hospitalization: A Cross-Sectional Study of Algeria  Do the vaccinated perform less distancing, mask wearing and hand hygiene? A test of the risk compensation hypothesis in a representative sample during the COVID-19 pandemic  Time trends in social contacts of individuals according to comorbidity and vaccination status, before and during the COVID-19 pandemic: repeated cross-sectional population-based surveys  Testing and Nonpharmaceutical Interventions for Prevention of SARS-CoV-2 in 20 US Overnight Camps in Summer 2021  Healthcare worker risk of COVID-19: A 20-month analysis of protective measures from vaccination and beyond  SARS-CoV-2 Transmission to Masked and Unmasked Close Contacts of University Students with COVID-19 - St. Louis, Missouri, January-May 2021  Mask usage, social distancing, racial, and gender correlates of COVID-19 vaccine intentions among adults in the US  Association between the COVID-19 Vaccine and Preventive Behaviors: Panel Data Analysis from Japan  Non-pharmaceutical interventions and risk of COVID-19 infection: survey of UK public from November 2020-May 2021  Impact of tiered measures on social contact and mixing patterns of in Italy during the second wave of COVID-19 |

**Table S4 Quality assessment**

| **NO.** | **Author** | **Study design** | | | | | | **Statistical methodology** | | | | |  | **Quality of reporting** | | | **Score**  **(total**  **=17)** |
| --- | --- | --- | --- | --- | --- | --- | --- | --- | --- | --- | --- | --- | --- | --- | --- | --- | --- |
|  |  | **Design** | **Source**  **of**  **data** | **Sample**  **size** | **Unbiased**  **inclusion of units** | **Level**  **of**  **inference** | **Prespecification**  **of ecological**  **units** | **Analytical**  **methodologies** | **Validity of**  **regression** | **Spatial**  **effects** | **Use of**  **covariates** | **Proper**  **adjustment**  **for**  **covariates** | **Internal**  **validity**  **of**  **the**  **methodology** | **Statement of**  **study**  **design** | **Justification**  **of**  **study**  **design** | **Discussion**  **cross-level**  **bias and**  **limitations** |  |
| 1 | Bollyky et al. | 1 | 1 | 2 | 1 | 1 | 1 | 1 | 1 | 1 | 1 | 1 | 1 | 1 | 1 | 1 | 16 |
| 2 | Ge et al.(1) | 1 | 1 | 1 | 1 | 1 | 1 | 2 | 1 | 1 | 1 | 1 | 1 | 1 | 1 | 1 | 16 |
| 3 | Paireau et al. | 1 | 1 | 2 | 1 | 1 | 1 | 2 | 1 | 1 | 1 | 1 | 1 | 1 | 0 | 0 | 15 |
| 4 | Ertem et al. | 1 | 1 | 2 | 1 | 1 | 1 | 1 | 1 | 1 | 1 | 1 | 0 | 1 | 1 | 1 | 15 |
| 5 | Zhou et al. | 1 | 1 | 1 | 1 | 1 | 1 | 2 | 1 | 1 | 1 | 1 | 1 | 1 | 0 | 1 | 15 |
| 6 | Huy et al. | 1 | 1 | 1 | 1 | 1 | 1 | 2 | 1 | 1 | 1 | 1 | 1 | 1 | 0 | 1 | 15 |
| 7 | Ge et al.(2) | 1 | 1 | 1 | 1 | 1 | 1 | 2 | 1 | 1 | 1 | 1 | 1 | 1 | 1 | 0 | 15 |
| 8 | Caixia Wang  et al. | 1 | 1 | 2 | 1 | 1 | 1 | 1 | 1 | 1 | 1 | 1 | 1 | 1 | 0 | 0 | 14 |
| 9 | Liang et al. | 1 | 1 | 1 | 1 | 1 | 1 | 2 | 1 | 1 | 1 | 1 | 1 | 1 | 0 | 0 | 14 |
| 10 | Jeonghyun  Shin  et al. | 1 | 1 | 2 | 1 | 1 | 1 | 1 | 1 | 0 | 1 | 1 | 0 | 1 | 0 | 1 | 13 |
| 11 | Hale et al. | 1 | 1 | 0 | 1 | 1 | 1 | 1 | 1 | 1 | 1 | 1 | 1 | 1 | 0 | 1 | 13 |
| 12 | Li et al. | 1 | 1 | 0 | 1 | 1 | 1 | 2 | 0 | 1 | 0 | 1 | 1 | 1 | 0 | 1 | 12 |
| 13 | Hongjian Wang  et al. | 1 | 1 | 2 | 1 | 1 | 1 | 2 | 0 | 1 | 0 | 0 | 0 | 0 | 0 | 1 | 11 |
| 14 | Sookhyun Kim  et al. | 1 | 1 | 2 | 1 | 1 | 1 | 1 | 0 | 0 | 0 | 0 | 1 | 0 | 0 | 1 | 10 |
| 15 | Kijin Kim et al. | 1 | 1 | 1 | 1 | 1 | 1 | 1 | 0 | 1 | 0 | 0 | 0 | 0 | 1 | 1 | 10 |
| 16 | Nesteruk | 1 | 1 | 1 | 1 | 1 | 1 | 1 | 0 | 1 | 0 | 1 | 0 | 1 | 0 | 0 | 10 |
| 17 | Nesteruk et al | 1 | 1 | 1 | 1 | 1 | 1 | 1 | 0 | 1 | 0 | 1 | 0 | 1 | 0 | 0 | 10 |

**Table S5 The characteristic of the included studies**

| **NPI Group** | **Author** | **Publication status, year** | **Geographical  scope** | **NPIs assessed** | **Outcome(s)** | **Source of NPIS data** | **Type of vaccination rate** | **Study time** | **Study time frame** | **Methods** | **Score**  **(total**  **=17)** |
| --- | --- | --- | --- | --- | --- | --- | --- | --- | --- | --- | --- |
| Composite indicator of NPIs | Ge et al. (1) | Published, 2022 | European,  33 countries | Stringency index^a^ | Rt | OxCGRT | Fully vaccine | Early and later stage | August 1, 2020 - October 25, 2021 | Bayesian inference model | 16 |
|  | Bollyky et al. | Published, 2023 | US,  52 states | Policy mandates^b^ | Cases,deaths | IHME COVID-19 database | Fully vaccine | Early | April 1,2020,to June 1,2021, | Regression analyses | 16 |
|  | Zhou et al. | Published, 2022 | European,  22 countries | Stringency index | Cases, deaths,  excess mortality | OxCGRT | Fully vaccine | Early stage | January 20, 2020-May 30, 2021 | Distribution lag model | 15 |
|  | Paireau et al. | Published, 2023 | France,  96 departments | Lockdown^c^ | Rt | Combination of governmental websites, press articles, and Wikipedia pages | First-dose | Early stage | From 9 March 2020 to 23 May 2021 | Log-linear mixed-effects model | 15 |
|  | Caixia Wang et al. | Published, 2023 | Worldwide,  176 countries | Stringency index | Deaths | OxCGRT | Fully vaccine | Omicron stage | June 15, 2021 to April 15, 2022 | Vector autoregression model | 14 |
|  | Li et al. | Published, 2022 | Worldwide,  8 countries | Combination of four NPIs^d^ | Rt | OxCGRT | First-dose | Later stage | January 21, 2020 -August 31, 2021 | Distributed lag non-linear model (DLNM) | 14 |
|  | Hale et al. | Published, 2021 | Worldwide,  10 countries | Stringency index | Deaths | OxCGRT | Unspecified | Early stage | January 1, 2020-March 11, 2021 | Ordinary Least Squares (OLS) regression | 13 |
|  | Hongjian Wang et al. | Published, 2023 | Worldwide,  176 countries | Stringency index | R0 | OxCGRT | Unspecified | Omicron stage | January 12, 2020 to May 14, 2022 | Dynamic Bayesian Network parameter learning | 11 |
|  | Kijin Kim et al. | Published, 2023 | Korea | Social distancing policy index^e^ | Cases | Refined social distancing policy index | Fully vaccine | Later stage | From February 2020 to October 14, 2021 | Vector autoregression model | 10 |

**Table S5 Continued**

| **NPI Group** | **Author** | **Publication status, year** | **Geographical  scope** | **NPIs assessed** | **Outcome(s)** | **Source of NPIS data** | **Type of vaccination rate** | **Study time** | **Study time frame** | **Methods** | **Score**  **(total**  **=17)** |
| --- | --- | --- | --- | --- | --- | --- | --- | --- | --- | --- | --- |
| Individual NPIs | Bollyky et al. | Published, 2023 | US, 52 states | Closures of bars, restaurants, gyms, and schools, facial covering and vaccine mandates, and stay-at-home orders and gathering restrictions | Cases,  deaths | IHME COVID-19 database | Fully vaccine | Early | April 1,2020,to June 1,2021, | Regression analyses | 16 |
|  | Huy et al. | Published, 2022 | Asina, 28 countries | School closure, workplace closure, public event canceling, public transport closure, stay at home requirements, restrictions on internal movement, international travel controls, public information campaign indicators ,testing policy, contact tracing, and facial covering | Growth rate | OxCGRT | First-dose | Early | 20 weeks of the pre- and post-vaccination period | Generalized linear mixed model (mGLMM) | 15 |
|  | Ge et al.(2) | Preprint, 2022 | Worldwide, 63 countries | School closures, workplace closures, gathering restrictions, movement restrictions, public transport closures, international travel restrictions, and facial coverings | Decay ratio | OxCGRT | First-dose | Early | 8 December 2020 - 25 March 2021 | Spatiotemporal Bayesian inference model | 15 |
|  | Ertem et al. | Published, 2023 | US, 2954 counties | Facial covering policies | Cases | The Yale State and Local COVI Drestriction database | First-dose | Early | April 4, 2020 to June 28, 2021 | Interrupted Time-Series Analysis | 15 |
|  | Paireau et al. | Published, 2023 | France, 96 departments | Curfews,school closures | Rt | Combination of governmental websites, press articles, and Wikipedia pages | First-dose | Early | March 9, 2020 to May 23, 2021 | Log-linear mixed-effects model | 15 |

**Table S5 continued**

| **NPI Group** | **Author** | **Publication status, year** | **Geographical  scope** | **NPIs assessed** | **Outcome(s)** | **Source of NPIS data** | **Type of vaccination rate** | **Study time** | **Study time frame** | **Methods** | **Score (total**  **=17)** |
| --- | --- | --- | --- | --- | --- | --- | --- | --- | --- | --- | --- |
| Individual NPIs | Liang et al. | Published, 2021 | Worldwide,  137 countries | School closures, workplace closures, cancellation of public events, restrictions on gathering size, requirements to stay-athome, and restrictions on international trave,public information campaigns, testing policy, contact tracing, face covering | Case doubling time | OxCGRT | First-dose | Early | January 1, 2020 - June 13,2021 | A random-effect growth-curve model with nonstandard interrupted time series analysis | 14 |
|  | Jeonghyun Shin et al. | Preprint,  2023 | India | Testing(Testing ratio) | Cases | Our World in Data | Fully vaccine | Later | April 28, 2021 - November 24, 2021 | Regression analysis | 13 |
|  | Li et al. | Published, 2022 | Worldwide,  8 countries | School closure; workplace closure; restrictions on public events; restrictions on gatherings; closure of public transport; stay-at-home requirements ; restrictions on internal movement; and international travel controls. | Rt | OxCGRT | First-dose | Later | January 21, 2020 -August 31, 2021 | Distributed lag non-linear model (DLNM) | 12 |
|  | Hongjian Wang et al. | Published, 2023 | Worldwide, 176 countries | Testing | R0 | Our World in Data | Unspecified | Omicron | January 12, 2020 to May 14, 2022 | Dynamic Bayesian Network parameter learning | 11 |
|  | Sookhyun Kim et al. | Published, 2023 | European,  35 countries | Facial covering policies | Cases | OxCGRT | Fully vaccine | Later | June 20 - October 30, 2021 | Correlations，t-test | 10 |
|  | Nesteruk | Preprint,  2022 | Worldwide,(Japan, Ukraine, USA; Hong Kong China,; mainland China; and European and African countries | Testing(testing ratio) | Cases | Our World in Data | Fully vaccine | Later, Omicron | 2020, 2021, 2022 | Non-linear correlation | 10 |
|  | Nesteruk et al. | Published,  2022 | Worldwide: 44 European and 14 other countries and regions | Testing(testing ratio) | Cases | Our World in Data | Fully vaccine, Boosters vaccine | Omicron | September 1,2021-February 1 , 2022 | Linear and Non-linear Regressions | 10 |
| Notes: (1) a= stringency index included eight containment and closure, including school closure, workplace closure, cancel public events, restrictions on gathering size, close public transport, stay-at-home requirements, restrictions on internal movement, restrictions on international travel；b =a summary measure that captures a state’s use of physical distancing and mask mandates；c=a comprehensive measure captures the level of social distancing; d=the combination of school closure, workplace closure, restrictions on mass gatherings and stay-at-home requirements; e=restrictiveness of government containment and closure measures implemented; Rt=effective reproductive number; R0=basic reproduction number. (2) Ge et al., Bollyky et al., and Li et al. analyzed both the effectiveness of the composite indicator of NPIs and individual NPIs. | | | | | | | | | | | |
